# Psychological Outcomes of Surgery Trainees in the Era of COVID-19 at a Tertiary Care Hospital of Karachi, Pakistan: A Cross-Sectional Study

**DOI:** 10.1101/2022.01.27.22269982

**Authors:** Gulzar Lakhani, Mohammad Hamza Bajwa, Nida Zahid, Syed Ather Enam

**Affiliations:** Department of Surgery, Aga Khan University Hospital, Karachi, Pakistan

**Keywords:** Surgical training, COVID-19, Burnout, Quality of Life, Generalized anxiety disorder

## Abstract

**Objectives:** The COVID-19 pandemic has resulted in interruptions in training for surgical residents in particular. This has been compounded by burnout and mental health concerns among surgical trainees across the world. We aim to determine the impact of the COVID-19 pandemic on psychological outcomes of surgical trainees in a tertiary-care hospital in Pakistan.

**Setting:** A cross-sectional, online survey-based study was conducted at a private tertiary care hospital of Karachi, Pakistan.

**Participants:** All the surgery trainees including; residents, fellows and instructors acquiring training at a private tertiary care hospital of Karachi, Pakistan were included in the study.

**Outcome measures:** The participants were assessed for their perceptions, and concerns regarding the COVID-19 pandemic, impact on their quality of life and generalized anxiety disorder (GAD).

**Results:** A majority (85%) of surgical trainees were concerned regarding contracting COVID-19 infections. Residents were more concerned with fellow residents developing burnout and anxiety as compared to their own psychological concerns. A strong, significant positive correlation (r=0.66, p <0.001) was noted between negative impact on QoL scores and developing GAD in surgical residents. On multivariable analysis, significant associations with GAD remained for family system, and negative impact of COVID-19 on QoL. The increased working hours for junior trainees showed more likelihood of developing GAD as compared to senior trainees.

**Conclusion:** Our investigation on QoL and psychological outcomes of surgical residents showed significant rates of burnout and GAD, which were attributed to concerns with the COVID-19 pandemic. We showed the impact this had on surgical trainees’ perception regarding their training and concerns with infecting others. This calls for structural interventions to address mental health concerns and provide psychological and educational support to surgical residents.

**Strengths and Limitations of the study:**

- Validated measures were used for evaluating the outcomes
- The study results can be generalized to all the surgery resident trainees working at private tertiary care hospitals.
- The response rate of trainees was 50%, the authors attempted to mitigate this through regular follow-up emails and reminders for responses.

## Introduction

The COVID-19 outbreak has had a significant impact on hospital staffing policies and structures. As we soon move into the third year of this pandemic, reports from across the world have documented the urgent need for restructuring resident and attending duty schedules, with many trainees being pulled from their specialties to provide care in high-volume and emergency care areas such as ICUs and emergency departments ^1^. Staff redeployments occurred due to a need to reduce inpatient services for non-urgent cases and shift towards COVID-19 intensive departments ^2^. This planning for every wave of the pandemic has ultimately taken a toll on front-line healthcare workers responsible for day-to-day care of these patients.

Surge planning has ultimately impacted training experiences with a significant toll on surgical residents ^3^. Graduated training in residency is incumbent upon trainees getting the required number of hours and specific case volumes to develop skills. This has invariably been affected with reports showing volume reduction in many specialties ^4^. Surgical training is unique with regards to the operating learning curve that requires in-person, hands-on training, without which residents may end up feeling inadequately prepared for independent practice. This has been significantly hampered with necessary precautions for COVID requiring increased PPE and reducing the amount of trainee surgeons able to be present for surgeries, in order to minimalize exposure ^5^. Case variety has also been reduced due to the shift towards emergency cases, significantly impacting residents looking to specialize.

While simulated training and technology-based learning has helped maintain educational standards, this has added to the stress already faced by residents all over the world ^6^. Residents themselves have been significantly impacted in terms of their physical wellbeing and mental health. Studies have shown that rates of burnout and mental health issues have increased as the pandemic has progressed, owing in part to the interruption in structured residencies ^7^. This is again compounded by many front-line healthcare workers themselves falling ill to this disease. These factors, together with the fear of being contagious and infecting others, could increase the possibility of psychological issues among them ^8^. Burnout and moral injury faced by healthcare workers in this pandemic have been seen across many countries with high levels of burnout being reported ^9^.

While ensuring we are doing everything possible for our patients, it is important to address systemic issues contributing to mental health concerns in residents. Currently, there is a paucity of data on the psychological health and quality of life (QoL) of surgical residents in Pakistan during the COVID-19 pandemic. Thus, we intend to evaluate surgical residents’ psychological health, perceptions and concerns regarding their training and the impact made by the COVID-19 pandemic.

## Methodology

### Study Design/ Site

It was a Cross sectional study. The surgery trainees were recruited from Department of Surgery, Aga Khan University Karachi, Pakistan.

### Study Participants

All the surgery trainees including; residents, fellows and instructors acquiring training at Aga Khan University Karachi, Pakistan were included in the study.

### Sampling Strategy/ Sample Size

We included all the surgery Trainees in our study. The Department of surgery has 10 sections and each section comprises of surgery trainees that includes; residents, fellows and instructors. The total number of residents, fellows and instructors are approximately 127. Following are the number of trainees in various specialties; Residents (22 General surgery, 19 Orthopedics, 10 Neurosurgery, 4 Plastic Surgery, 10 Urology, 5 Pediatric Surgery, 16 Dental Surgery, 9 Head and Neck Surgery, 5 Ophthalmology and 7 Cardiac Surgery); Fellows (2 General Surgery, 1 Breast Surgery, 1 Head and Neck Surgery and 2 Vascular Surgery); Instructors (2 General Surgery, 2 Orthopedics, 1 Neurosurgery, 1 Dental Surgery and 1 Cardiac Surgery). However, we were able to collect data of 59 trainees.

### Eligibility Criteria

Inclusion Criteria

1. All the surgery trainees acquiring training at Department of Surgery Aga Khan University Hospital Karachi, Pakistan
2. Those participants who filled the online questionnaire Exclusion
3. All those participants who did not completed the form

### Data Collection

The data was collected on online survey monkey form. The form is divided into 4 sections. Section A comprises of demographics information such as, age, gender, place of residence, ethnicity, marital status, type of family, household family members. Information will also be taken on type of specialty and year of training. Section B comprises of resident perceptions of risk for contracting covid-19 and concerns surrounding the novel coronavirus disease 2019 pandemic.^10^ Section C is the COVID-19–Impact on Quality of Life (COV19-QoL) scale v1.5^11^, which will be used to determine the impact of covid on QoL of the surgery trainees. It is a 6 items tool on a Likert scale of 1-5. A high score determines a lower QoL. Section D is generalized anxiety disorder 7-item (gad-7) scale. ^12^ It is on a Likert scale of 0-3.

### Ethical Considerations

The Ethical exemption was obtained from Aga Khan University, ethics review committee (AKU-ERC) since there was no interaction with the participants and information was collected via an online Google form without identities. All the information obtained from the surgery trainee was kept confidential. The online form and the data base is password protected. No personal identifier was disclosed. The participation of the trainee in this survey was voluntary.

### Plan of Analysis

Data was analyzed on STATA version 12. Descriptive data for quantitative variables were reported as mean ± SD/ median (IQR) and for qualitative variables as frequency and percentages. Pearson correlation coefficient (r) was reported by correlation analysis to assess correlation between GAD and QoL. Unadjusted and adjusted beta coefficient with their 95% by using linear regression analysis was reported to assess the factors associated with GAD among surgery trainee. All plausible interactions were assessed. A p value of < 0.2 was considered as significant for univariate analysis and a p value of < 0.05 will be considered significant for multivariable analysis.

### Patient and Public Involvement

This was a cross-sectional study design and the participants information regarding their socio-demographic factors, anxiety and QoL, was assessed online via Google form. The study findings will be disseminated to different stakeholders, such as healthcare professionals, rehabilitation experts, and psychologists through: publications at local, national and international journals, presentations at conferences and workshops and through research briefs.

## Results

### Participant Characteristics

A total of 59 out of 127 (46%) participants responded to the online survey. The mean age of the participants was 29.05 ± 2.26 years and 55.9% were males. 96.6% of participants were residents of Karachi. The mother tongue of 30.8% participant was Urdu. Most of the participants (69.5%) were unmarried and half of them lived in nuclear families. 70.6% of participants had spouses who were working. About 55.6% of the married participants had children. 74.6% had family members above 60 years old while 49.2% had children in their households. **(Table 1)**

**Table 1:**
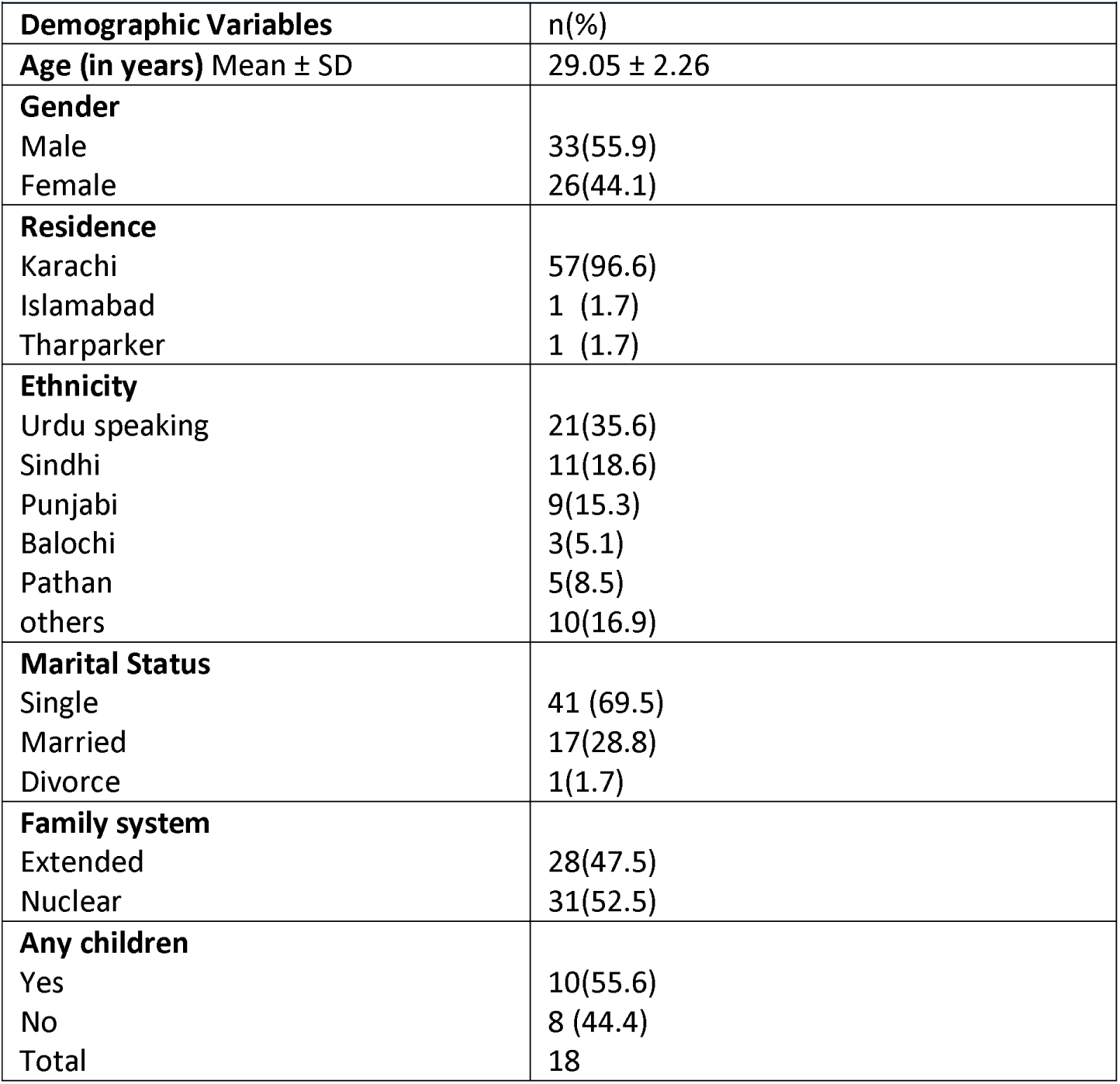
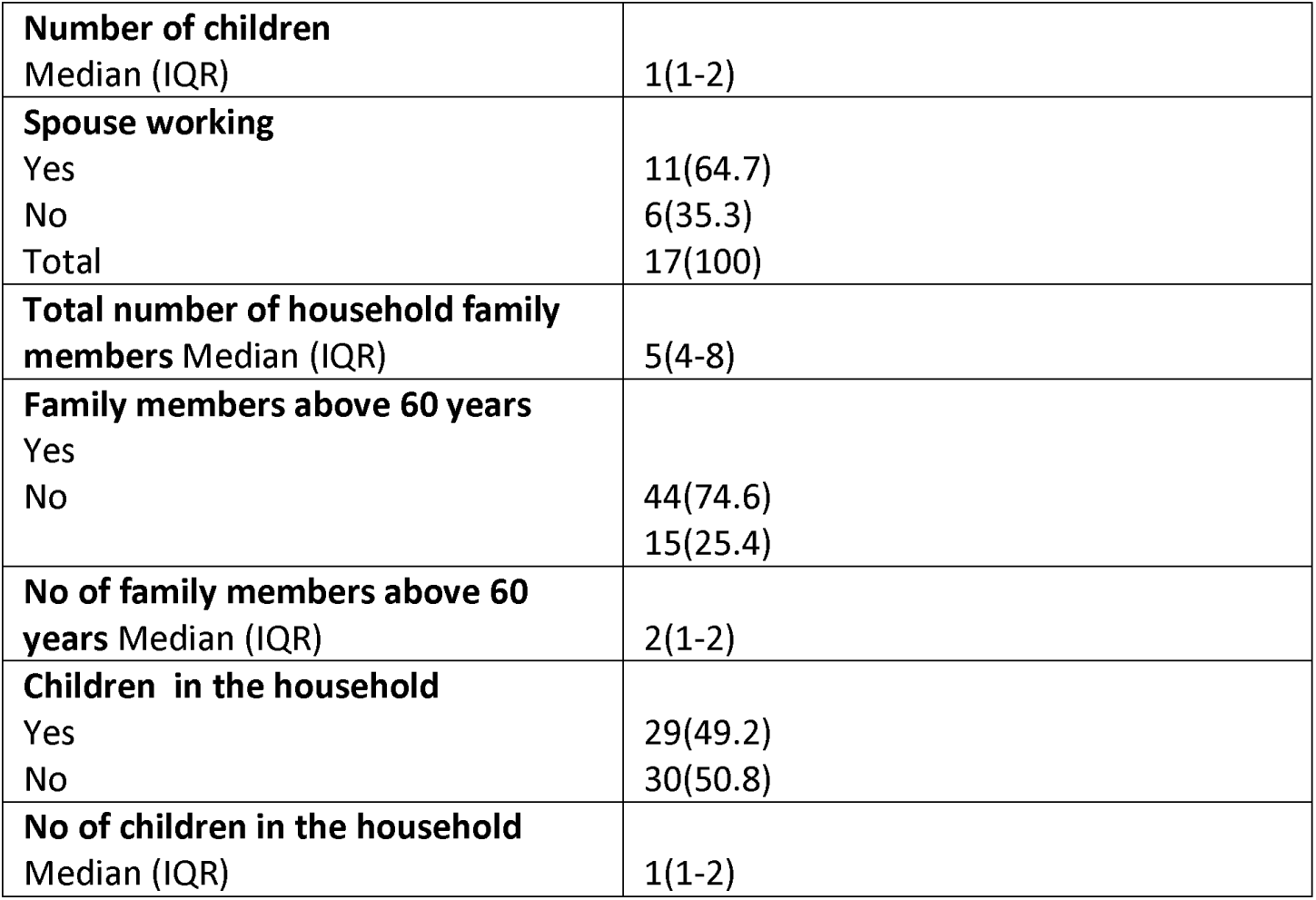
Demographics of the surgery trainees (N=59)

### Training status and working hours of the Surgery Trainees

The **table 2** shows the training status of the participants. About 94.9% of the participants were residents. A higher proportion (50%) of the participants were in their early phase of their residency i.e (year 1 and 2). The trainees worked for 80 (40-98) hours per week. A higher proportion of trainees (25.4%) were from dentistry followed by general surgery 22%, neurosurgery 13.6%, orthopedic 11.9% etc **(Figure 1)**

**Table 2:** Training status and working hours of the Surgery Trainees (N=59)

| **Table 2: Training status and working hours of the Surgery Trainees (N=59)** |  |
| --- | --- |
| Training status | n (%) |
| **Type of Trainee**<br>Residents<br>Fellows | 56 (94.9)<br>3(5.1) |
| **Year of Training**<br>Year 1<br>Year 2<br>Year 3<br>Year 4<br>Year 5<br>Year 6 | 15 (25.4)<br>16(27.1)<br>11(18.6)<br>6(10.2)<br>7(11.9)<br>4(6.8) |
| **Number of work days per week on duty** Median (IQR) | 6(6) |
| **Number of work hours per week on duty** Median (IQR) | 80(40-98) |

**Figure 1:**
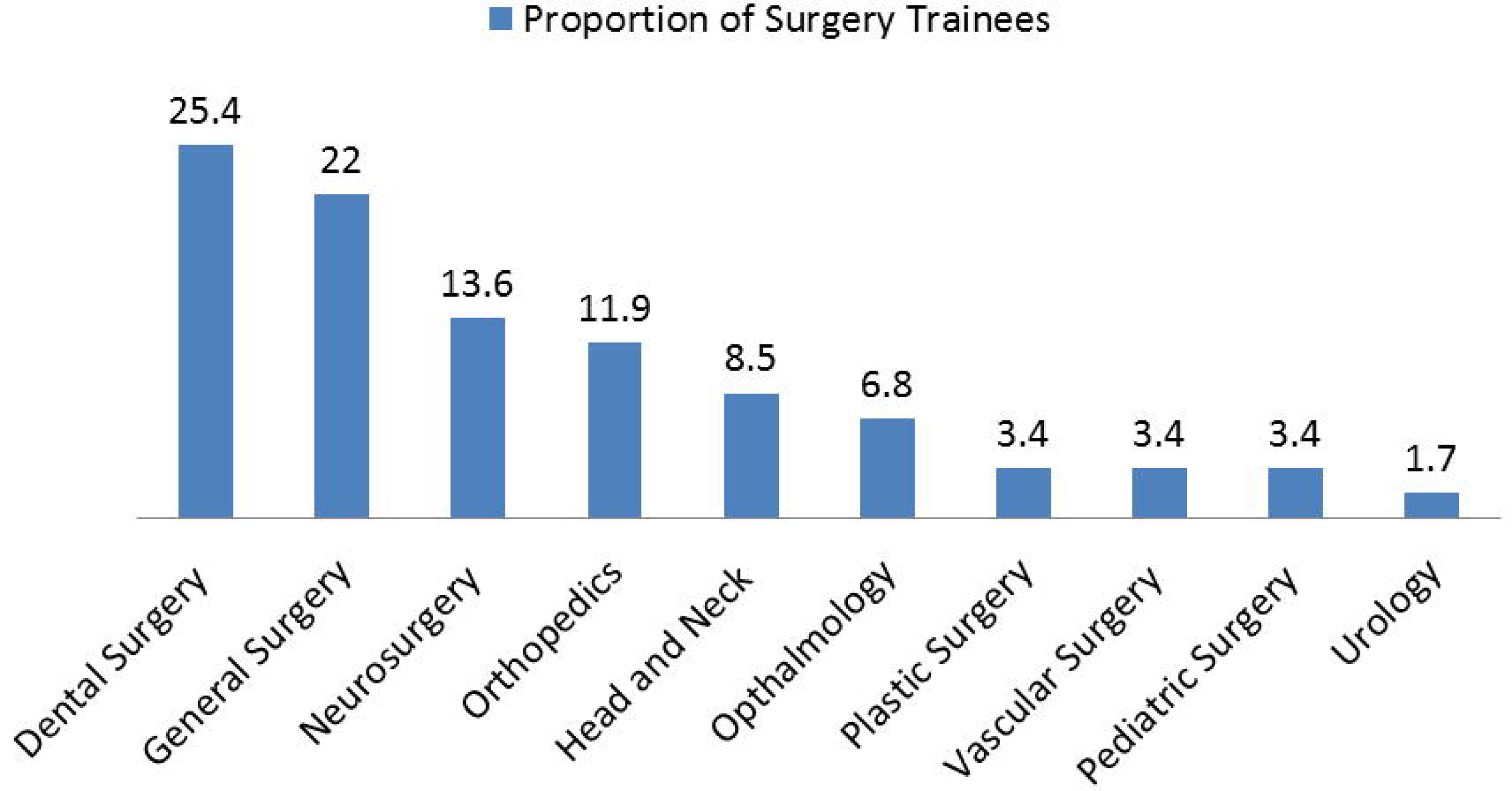
Proportion of Trainees in different Surgical Specialties.

### Perception and concerns of the Surgery Trainees

Majority of the participants’ (85%) were highly concerned about risk of contracting Covid **(Figure 2a)**. A higher proportion (67%) of the participants were moderately to extremely concerned about their co-residents burn out and anxiety as compared to their own anxiety and burnout (52.5%). 66% of the participants (73%) were moderately to extremely concerned about contracting Covid. Moreover, majority of the participants were moderately to extremely concerned about impact of Covid on their clinical and surgical education. A higher proportion of the participants (80%) were moderately to extremely concerned about transmitting Covid to family and friends as compared to transmitting it to their patients (51%). Majority of the participants (35%) were not concerned about shortage of PPE. **(Figure 2b)**

**Figure 2a:**
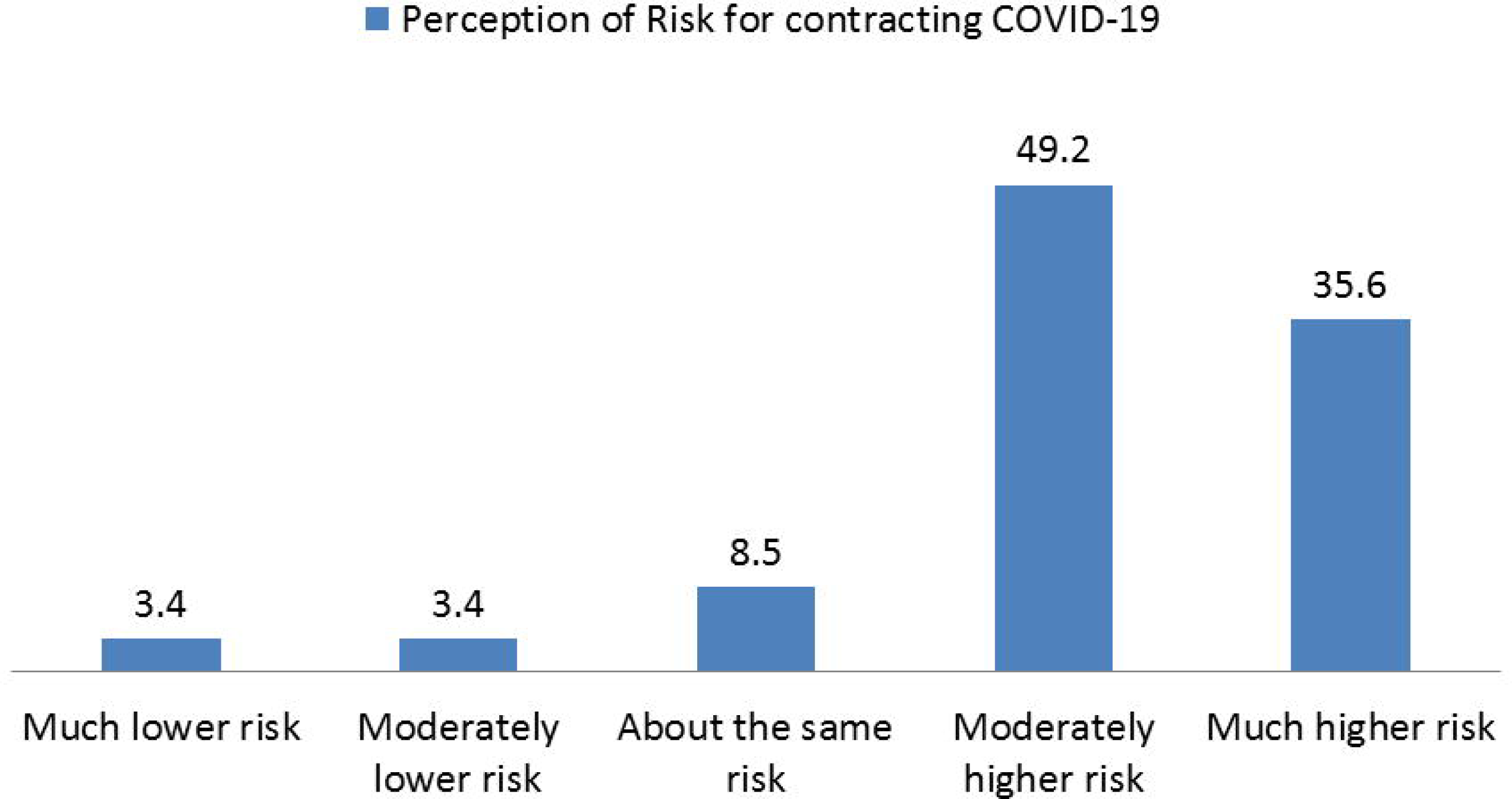
Perception and concerns of the Surgery Trainees surrounding the COVID 19 Pandemic.

**Figure 2b:**
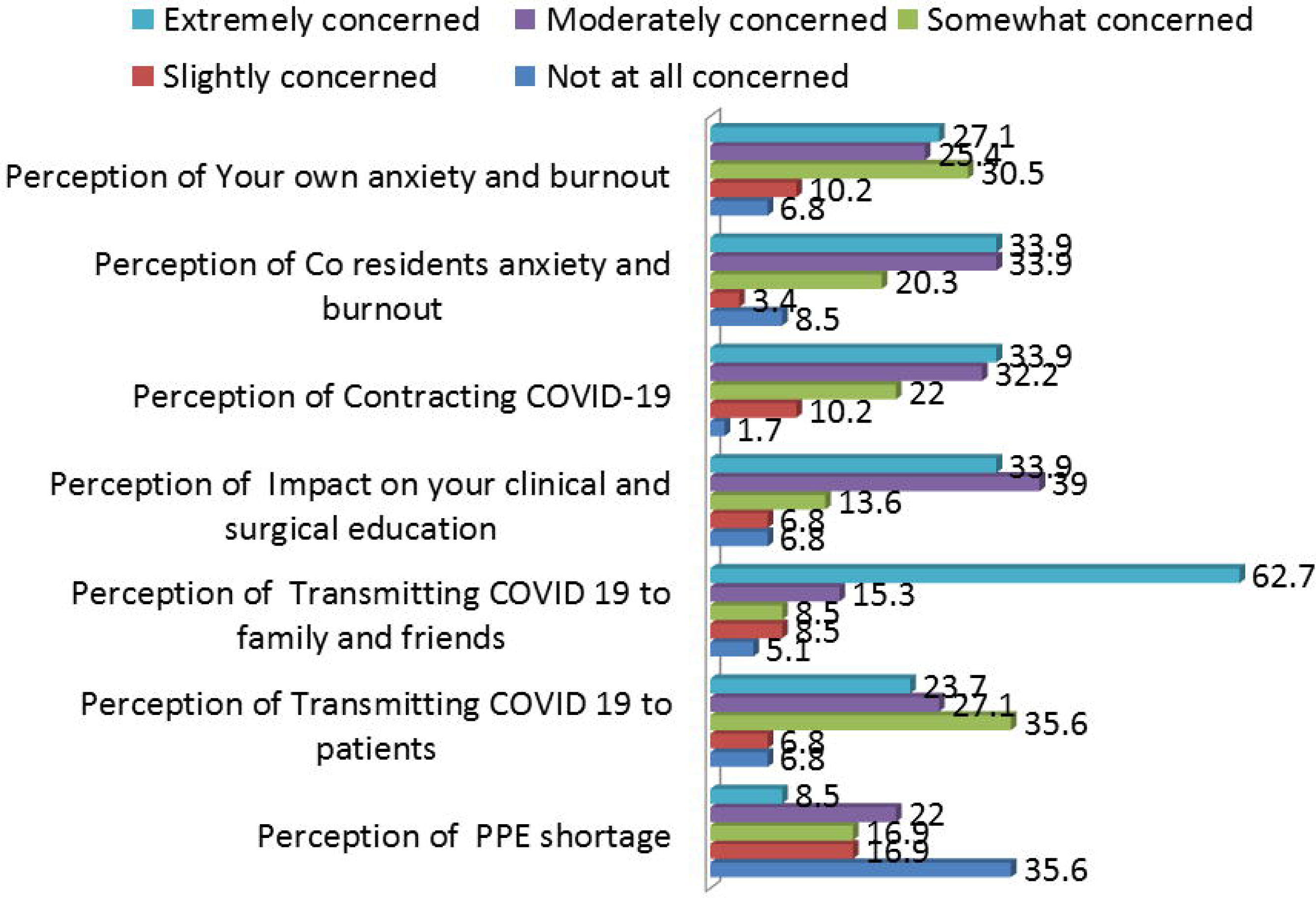
Perception of trainees of Risks and Concerns regarding COVID 19.

### Mental Health of Surgery Trainees

**Table 3** depicts the mental health of the surgery trainees. We observed that Covid had high negative impact on the QoL of 47.5% participants. Moreover, 25% of the participants had clinically significant general anxiety disorder.

**Table 3:** Mental Health of Surgery Trainees

| **Table 3: Mental Health of Surgery Trainees** |  |
| --- | --- |
| **Impact of Covid on quality of life (QoL) of Trainees** |  |
| **Overall Impact of COVID 19 on QoL** |  |
| Low negative Impact ($\leq 3$) | 31(52.5) |
| High negative impact ($>3$) | 28(47.5) |
| Negative impact of Covid on QoL Score (Mean ± SE) | 3.05±0.12 |
| I think my quality of life is lower than before (Mean ± SE) | 3.07 ±0.14 |
| I think my mental health has deteriorated(Mean ± SE) | 2.83±0.16 |
| I think my physical health may deteriorate(Mean ± SE) | 3.36 ±0.13 |
| I feel more tense than before(Mean ± SE) | 3.08 ± 0.16 |
| I feel more depressed than before(Mean ± SE) | 2.91± 0.16 |
| **Generalized Anxiety Disorder (GAD) among trainees** |  |
| **Total GAD score**<br>(Mean ± SE) | 6.74 ±0.76 |
| **GAD** |  |
| No Anxiety | 30(50.8) |
| Mild Anxiety | 14(23.7) |
| Moderate Anxiety | 10(16.9) |
| Severe Anxiety | 05(8.5) |
| **GAD** |  |
| Clinically insignificant (< 3) | 44(74.6) |
| Clinically significant (≥3) | 15(25.4) |

### Correlation of Generalized Anxiety Disorder And Impact of Covid on quality of Life

**Table 4** shows correlation between negative impact of COVID 19 on QoL and GAD. There was a strong significant positive correlation (r=0.66, p <0.001) between negative impact on QoL scores and GAD. We observed a strong significant positive correlation between the following domains; feel more tense than before and GAD (r=0.62, p=<0.001) and feel more depress than before and GAD (r=0.71, p <0.001). A moderate significant positive correlation between mental health has deteriorated than before and GAD (r=0.57, p <0.001) and quality of life is lower than before and GAD (r=0.48, p <0.001). However there was no significant correlation between ‘physical health may deteriorate’ and GAD at p < 0.05.

**Table 4.**
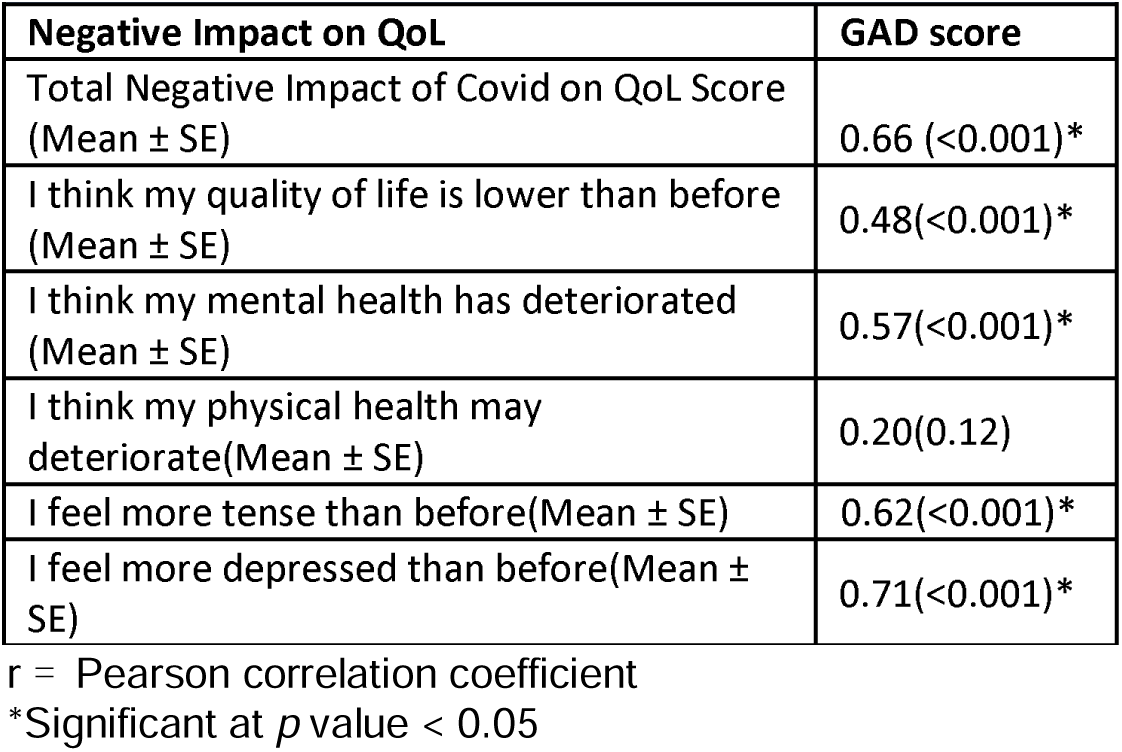
Correlation of Generalized Anxiety Disorder And Impact of COVID 19 on quality of Life among surgery trainees

| **Table 4. Correlation of Generalized Anxiety Disorder And Impact of Covid on quality of Life among surgery trainees** |  |
| --- | --- |
| **Negative Impact on QoL** | **GAD score** |
| Total Negative Impact of Covid on QoL Score (Mean ± SE) | 0.66 (<0.001)* |
| I think my quality of life is lower than before (Mean ± SE) | 0.48(<0.001)* |
| I think my mental health has deteriorated (Mean ± SE) | 0.57(<0.001)* |
| I think my physical health may deteriorate (Mean ± SE) | 0.20(0.12) |
| I feel more tense than before (Mean ± SE) | 0.62(<0.001)* |
| I feel more depressed than before (Mean ± SE) | 0.71(<0.001)* |

$r$ = Pearson correlation coefficient
\*Significant at $p$ value < 0.05

### Univariate and multivariable linear regression with generalized anxiety disorder as dependent variable

On Univariate analysis we observed that GAD was 3 units significantly higher among females as compared to males. With every 1 unit increase in age GAD was decreased by 0.63 units. Participants living in nuclear family had significantly higher GAD compared to those living in extended family. Participants whose spouses were not working had a significantly lower GAD compared to those whose spouses were working. Participants with more family member in their household had significantly lower GAD (ß=-0.34 (−0.84, 0.15). Similarly, participants with children in their household had 2 units significantly lower GAD as compared to those with no children. Participants with high negative impact of COVID-19 on their QoL had 7 units significantly higher GAD as compared to those with low negative impact on QoL. Year of residency and work hours also had a significant association with GAD at p <0.2. **(Table 5)**

**Table 5.**
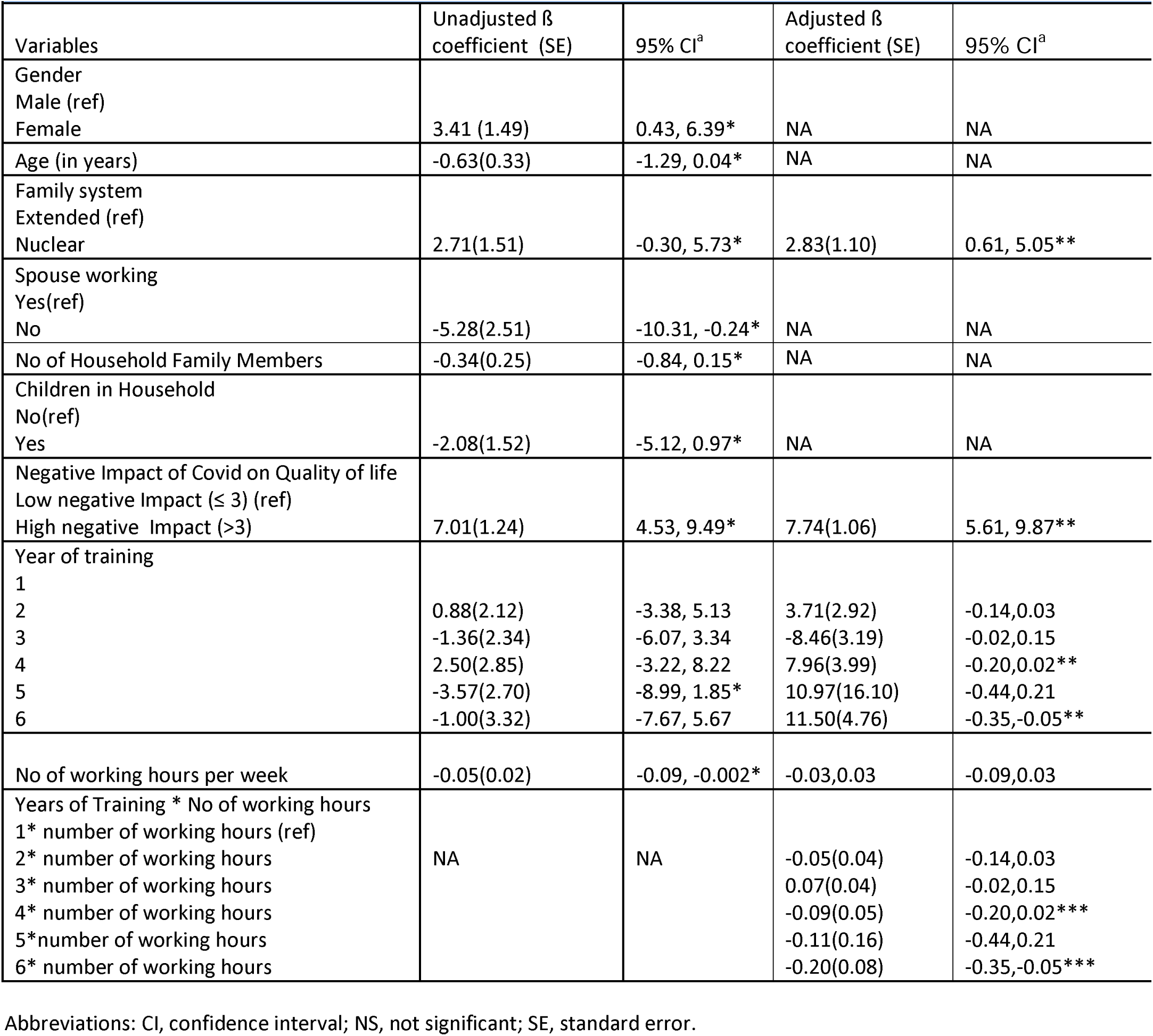
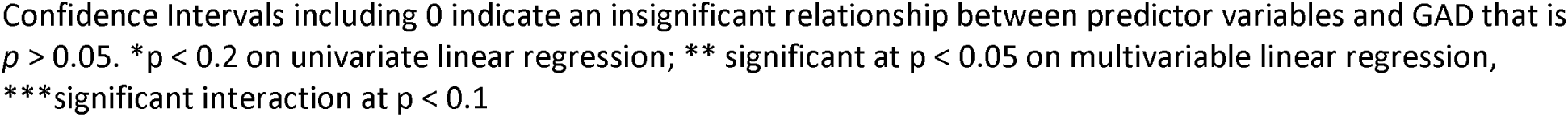
Univariate and multivariable linear regression with Generalized Anxiety Disorder as dependent variable

|  |  |  |  |  |
| --- | --- | --- | --- | --- |
| Variables | Unadjusted $\beta$ coefficient (SE) | 95% CI<sup>a</sup> | Adjusted $\beta$ coefficient (SE) | 95% CI<sup>a</sup> |
| Gender |  |  |  |  |
| Male (ref) |  |  |  |  |
| Female | 3.41 (1.49) | 0.43, 6.39* | NA | NA |
| Age (in years) | -0.63(0.33) | -1.29, 0.04* | NA | NA |
| Family system |  |  |  |  |
| Extended (ref) |  |  |  |  |
| Nuclear | 2.71(1.51) | -0.30, 5.73* | 2.83(1.10) | 0.61, 5.05** |
| Spouse working |  |  |  |  |
| Yes(ref) |  |  |  |  |
| No | -5.28(2.51) | -10.31, -0.24* | NA | NA |
| No of Household Family Members | -0.34(0.25) | -0.84, 0.15* | NA | NA |
| Children in Household |  |  |  |  |
| No(ref) |  |  |  |  |
| Yes | -2.08(1.52) | -5.12, 0.97* | NA | NA |
| Negative Impact of Covid on Quality of life |  |  |  |  |
| Low negative Impact ($\leq 3$) (ref) | | | | |
| High negative Impact ($>3$) | 7.01(1.24) | 4.53, 9.49* | 7.74(1.06) | 5.61, 9.87** |
| Year of training |  |  |  |  |
| 1 |  |  |  |  |
| 2 | 0.88(2.12) | -3.38, 5.13 | 3.71(2.92) | -0.14, 0.03 |
| 3 | -1.36(2.34) | -6.07, 3.34 | -8.46(3.19) | -0.02, 0.15 |
| 4 | 2.50(2.85) | -3.22, 8.22 | 7.96(3.99) | -0.20, 0.02** |
| 5 | -3.57(2.70) | -8.99, 1.85* | 10.97(16.10) | -0.44, 0.21 |
| 6 | -1.00(3.32) | -7.67, 5.67 | 11.50(4.76) | -0.35, -0.05** |
| No of working hours per week | -0.05(0.02) | -0.09, -0.002* | -0.03, 0.03 | -0.09, 0.03 |
| Years of Training * No of working hours |  |  |  |  |
| 1* number of working hours (ref) |  |  |  |  |
| 2* number of working hours | NA | NA | -0.05(0.04) | -0.14, 0.03 |
| 3* number of working hours |  |  | 0.07(0.04) | -0.02, 0.15 |
| 4* number of working hours |  |  | -0.09(0.05) | -0.20, 0.02*** |
| 5* number of working hours |  |  | -0.11(0.16) | -0.44, 0.21 |
| 6* number of working hours |  |  | -0.20(0.08) | -0.35, -0.05*** |

Abbreviations: CI, confidence interval; NS, not significant; SE, standard error.
Confidence Intervals including 0 indicate an insignificant relationship between predictor variables and GAD that is $p > 0.05$ . \* $p < 0.2$ on univariate linear regression; \*\* significant at $p < 0.05$ on multivariable linear regression, \*\*\*significant interaction at $p < 0.1$

On multivariable analysis we observed a significant association of family system and negative impact of COVID-19 on QoL. There was also a significant interaction between years of residency and working hours per week. We observed that GAD was 2.8 units significantly higher in participants living in nuclear family as compared to those living in extended family. Moreover, participants with high negative impact of COVID-19 on their QoL had 7.7 units significantly higher GAD as compared to those with low negative impact on QoL. There was significant interaction between year of training and working hours. With every one hour increase in work the year 1 trainees were 0.2 units more likely to have GAD compared to year 6 trainees. Similarly, with every one hour increase in work the year 1 trainees were 0.09 units more likely to have GAD compared to year 4 trainees. However, GAD of year 1 trainees was not significantly different compared to years 2, 3 and 5 trainees. **(Table 5)**

## Discussion

Overall, our study showed significant deterioration in the quality of life of surgical residents secondary to the COVID-19 pandemic. As can be seen from our assessment of perceptions of residents, the cause is both due to risks associated with infection to those nearest to them i.e. colleagues and family, as well as the significant impact made on training. Particularly in terms of psychological health, a portion of residents had symptoms on par with diagnosed GAD. Through our QoL questionnaire, we were able to see a significant correlation of the negative impact on QoL with GAD. Analyzing the risk factors significant for developing GAD, female residents, residents living in nuclear families, residents with working spouses, and higher-level surgical residents were most likely to develop GAD during the pandemic.

Interestingly, we observed that increased work hours for PGY-1 trainees showed more likelihood of developing GAD in comparison with PGY-4 and PGY-6 trainees. We did not see any significantly different likelihood of developing GAD between PGY-1 residents and PGY-2, 3, and 5 residents.

The impact of the COVID-19 pandemic on surgical trainees in Pakistan has been investigated by only one other paper; the authors reported statistically significant reduction in burnout among surgical residents according to the Maslach Burnout inventory ^13^. This was attributed to reduced average number of working hours per week. However, similar to our study, the cohort of trainees reported similar adverse effects on surgical hands-on exposure and fears of transmitting COVID 19 to family members. As this study was conducted a year before our current report, we can understand our findings are an update to the current status of mental health in surgical trainees in Pakistan, with a better evaluation of the chronic effects this has had on psychological and QoL outcomes.

To the best of our knowledge, this study is the first investigation into the QoL impact of the COVID-19 pandemic in Pakistan.These findings are comparable to QoL surveys done in other countries as well. A recent study of surgical trainees in the UK redeployed to other specialties showed that over 50% of trainees felt that the interruptions due to the COVID-19 pandemic had a negative impact on their mental health ^14^. Participants in the study reported lack of operating and implications on skills development, burnout, health of family and colleagues, and isolation were main contributors to negative mental health outcomes. This has been seen within our cohort as well. Overall, trainees have reported worse mental health outcomes and increased burnout compared to pre-pandemic rates ^15^.

In our study we observed that senior residents were less anxious as compared to junior residents. Similar to our findings, a study reported that PGY1 medical residents reported a chronic sleep deprivation, depression, and burnout. ^16^ Another study of residents suggested that senior residents (PGY3) had the highest QoL compared to junior residents (PGY1 and 2). ^17^

Our study’s findings also showed that females had higher generalized disorder scores as compared to males. Our study’s findings were consistent to that also found that female residents had more work life balance dissatisfaction and emotional exhaustion. ^17^ The plausible explanation of this could be that females have additional demands outside their work place and family stresses.^18^

Our findings also show that trainees who live in a nuclear family system have a higher rate of generalized anxiety disorder than those who live in extended families. Our findings were similar to those of a study conducted on students in Bangladesh, that found that students who live alone or with fewer family members are more likely to experience anxiety during COVID 19.^19^This could be because people who live in extended families can share their worries, stresses, and responsibilities with other family members amidst COVID 19 pandemic.

Our study is limited by the response rate of trainees we observed; the authors attempted to mitigate this through regular follow-up emails and reminders for responses. Various response rates for email/web-based surveys have been reported in the literature, ranging from 13% ^20^ to 80%. ^21 22^ It’s vital to keep in mind how frequently residents and staff physicians are receive such surveys. Since these survey requests add to already heavy workloads, residents and staff physicians are unlikely to complete them unless there is a incentive or it is an essential requirement. Additionally, our experience is of a private-sector, tertiary care hospital. This may be different to the impact of COVID-19 on surgical trainees in public-sector hospitals, where significant constraints exist in managing COVID-19 patients.

It is vital that we address these concerns faced by surgical trainees. The impact of burnout and deterioration in QoL can lead to reduced motivation, errors made in patient care, and chronic fatigue in trainees than can further the vicious cycle ^23^. Creating programs to facilitate residents’ education through virtual platforms and develop systems of support can ameliorate residents’ concerns ^24^. Educational institutions have a particular responsibility to maintain training program standards. Centers that invest in surgical skills simulation training, virtual conferences, and regulating re-allocation to ensure adequate participation of trainees in core competency development have shown positive results in resident education ^25^. It is through ensuring the wellbeing of surgical residents that we can promise a better return to workflow once restrictions are no longer in place.

## Conclusion

The investigation into QoL and psychological outcomes of surgical residents showed a significant rate of burnout and GAD, which were significantly associated with the COVID 19 pandemic. Our study also showcases the concerns faced by surgical residents through their training and interactions with COVID-19 patients. Significant risk factors for developing GAD in the pandemic were residents without extended family support, high negative impact of COVID on QoL, and higher year of training. We recommend structural improvements to address mental health concerns as well as educational concerns of residents with regards to training standards.

## Data Availability

Data will be available upon request from the corresponding author

## Contributors

GL contributed to conception of the study, manuscript drafting and reviewing. MHB was responsible for manuscript writing and reviewing the paper. NZ helped in data analysis, manuscript writing and reviewing of the manuscript. SAE contributed to conception of the study, manuscript drafting and reviewing the paper. All authors saw and approved the final version of manuscript.

## Participants consent for publication

Not required.

## Ethics approval

Study protocol is exempted by Aga khan university ethical review committee with ERC # 2020-5092-11410.

## Provenance and peer review

Not commissioned; externally peer reviewed

## Data availability statement

Data will be available upon request from the corresponding author

### List of Abbreviations

PGY: Post graduate year
GAD: Generalized anxiety disorder
QoL: Quality of Life
AKU: Aga Khan University
ERC: Ethical Review Committee
SD: Standard Deviation
IQR: Inter quartile range

